# Development And Performance Evaluation of A Rapid In-House ELISA for Retrospective Serosurveillance of SARS-CoV-2

**DOI:** 10.1101/2020.12.10.20244350

**Authors:** Bijon Kumar Sil, Mumtarin Jannat Oishee, Md. Ahsanul Haq, Nowshin Jahan, Tamanna Ali, Shahad Saif Khandker, Eiry Kobatake, Masayasu Mie, Mohib Ullah Khondoker, Mohd. Raeed Jamiruddin, Nihad Adnan

**Affiliations:** Gonoshasthaya-RNA Molecular Diagnostic & Research Center, Dhanmondi, Dhaka-1205; Department of Microbiology, Gono Bishwabidyalay, Savar, Dhaka-1344, Bangladesh; Department of Life Science and Technology, Tokyo Institute of Technology; Gonoshasthaya Samaj Vittik Medical College, Savar, Dhaka-1344, Bangladesh; Department of Pharmacy, BRAC University, Dhaka-1212, Bangladesh; Department of Microbiology, Jahangirnagar University, Savar, Dhaka-1342, Bangladesh

**Keywords:** SARS-CoV-2, COVID-19, serosurveillance, ELISA, nucleocapsid

## Abstract

**Background:** In the ongoing pandemic situation of COVID-19, serological tests can complement the molecular diagnostic methods, and can be one of the important tools of sero-surveillance and vaccine evaluation.

**Aim:** To develop and evaluate a rapid SARS-CoV-2 specific ELISA for detection of anti-SARS-CoV2 IgG from patient’s biological samples.

**Methods:** In order to develop the ELISA, three panels of samples (n=184) have been used: panel 1 (n=19) and panel 2 (n=60) were collected from RT-PCR positive patients within 14 and after 14 days of onset of clinical symptoms respectively, whereas panel 3 consisted of negative samples (n=105) collected either from healthy donors or pre-pandemic dengue patients. As a capturing agent full-length SARS-CoV2 specific recombinant nucleocapsid was immobilized. Commercial SARS-CoV2 IgG kit based on chemiluminescent assay was used for the selection of samples and optimization of the assay. The threshold cut-off point, inter-assay and intra-assay variations were determined. The total assay time for this in-house ELISA was set for 30 minutes.

**Results:** The assay time was set at a total of 30 minutes with the sensitivity of 84% (95% confidence interval, CI, 60.4%, 96.6%) and 98% (95% CI, 91.1%, 100.0%), for panel 1 and 2 respectively, with over all 94.9% sensitivity (95% CI 87.5%, 98.6%). Moreover, the clinical specificity is 97.1% (95% CI, 91.9%, 99.4%) with no cross reaction with dengue sample. The overall positive and negative predictive values are 96.2% (95% CI 89.2%, 99.2%) and 96.2% (95% CI, 90.6% 99.0%) respectively. In-house ELISA demonstrated 100% positive and negative percent agreement with ROCHE (Elecsys; Anti-SARS-CoV-2), with a Cohen’s kappa value of 1.00 (very strong agreement), while comparing 13 positive and 17 negative confirmed cases.

**Conclusion:** The assay is rapid and can be applied as one of the early and retrospective sero-monitoring tools in all over the affected areas.

## Introduction

The current situation of the world is all about the war between the visible and invisible. Life has to adopt a new normality due to the advent of an acute respiratory disease, COVID-19 (1). The disease, emerged in December 2019 in China, has been evolved as a public health threat due to its global spread (2), morbidity and mortality rate. The etiological agent of this is SARS-CoV-2, a positive strand RNA virus, belonging to the beta-coronavirus family (3). The disease presents an unprecedented spectrum in clinical manifestations ranging from asymptomatic, mild or quasi-common-cold symptoms to severe complications requiring immediate medical intervention (4-6). Droplet, airborne, orofecal and fomite transmission of this virus as well as direct contact with symptomatic and asymptomatic individuals contribute to the rampant spread of the disease (5, 7). The havoc brought down by the pandemic demands early diagnosis during the acute phase of infection. Viral RNA detection by real time RT-PCR is the gold standard for early diagnosis (8). This immediate step identifies acute illness facilitating disease management and restricting rapid spread to some extent. However, collection of samples from suitable sites, sample transportation, the constraints of efficient and trained personnel as well as well-equipped facilities and increased false negative results of RT-PCR at later phase of infection disqualifies its sole implementation in the field of SARS-CoV2 diagnostics (9, 10). Although other molecular based methods such as isothermal amplification techniques or CRISPR-based technology are implemented and suggested, are yet to be well-practised considering the cost-effectiveness (11, 12).

Though serological tests are not yet suggested for case detection by World Health Organization, in order to reveal the scenario of the prevalent and past episodes, serological assay has to be of prime importance (13, 14). Retrospective serosurveillance, not only enlightens with current immune status of the exposed individuals, but also facilitates therapeutic action by selecting convalescent plasma donor as well as to study the plausible outcome from the vaccine shoot focused on neutralizing antibody (15). Mass screening of a population is required to move towards relaxing COVID-19 restrictions. Decisions like ‘back to the work’ and school require timely seroprevalence study. All these aspects necessitate highly sensitive and specific immunoassay (16, 17).

The key structural proteins of SARS-CoV2 include Nuclecapsid protein (NCP), Spike (S), Envelop protein (E) and Membrane (M) protein. Of these the N and S proteins are highly immunogenic in nature commencing generation of IgM and IgG antibodies (18). These proteins are now exploited as suitable targets for developing several serological assays like Enzyme Linked Immunosorbent Assay (ELISA) (19).

The nucleoprotein of SARS-CoV-2 is highly immunogenic and detection of antibody against this protein is found to be more sensitive compared to Spike (S) or RBD (20, 21). This study characterizes an in-house ELISA targeting IgG antibody against full-length SARS-CoV-2 nucleocapsid protein (NCP). To our knowledge, it is the first indigenous ELISA that is rapid with thirty minutes incubation time and possessing higher sensitivity and specificity. Three panels comprising of a total of 184 samples have been considered for the development of this ELISA and a comparative study has been done with Elecsys Anti-SARS-CoV-2 which is based on chemiluminescent assay.

## Materials and methods

### Reagents

Recombinant nucleocapsid protein specific to SARS-CoV-2 was obtained from Sino Biologicals, China, and used as capturing agent. Goat anti-human IgG conjugated with HRP (Native Antigen, UK) was used to detect human IgG which formed an immune-complex with coated SARS-CoV-2 specific antigen. 3,3′,5,5′-Tetramethylbenzidine (TMB) (Dojindo Molecular Technologies, USA) was used as colour developer suitable for peroxidase substrate (Wako, Japan) while 1.5 M H_2_SO_4_ (Sigma-Aldrich, Germany) was used to stop the colour developed by TMB-peroxidase and read at 450 nm using ELISA plate reader (Thermo-Fisher Scientific, USA).

### Sample selection and panel composition

The assay was developed and evaluated using panels of serum samples as per the FDA guidance documents “Policy for Coronavirus Disease-2019 Tests During the Public Health Emergency (Revised)” and FDA recommendation. Three panels of serum samples comprising in total of 184 were collected from 134 individuals with their proper consent. Panel-1 comprises of 19 RT-PCR positive samples collected from fourteen COVID-19 patients within 2 weeks of their onset of clinical symptoms of infection, while the second panel consisted of sixty RT-PCR positive samples collected after 14 days of onset of symptoms from patients. Panel 3 included eighty one serum samples from healthy donors collected between April to June 2020, while 24 samples were collected from dengue positive patients before the outbreak of COVID-19. All the samples were stored in -80 °C until further analysis.

### Characterization of seropositive and seronegative samples by Commercial Kit

A total of thirty samples of which thirteen from COVID-19 RT-PCR positive individuals and seventeen from healthy donors were characterized using one of the Food and Drug Administration (FDA) EUA approved commercially available chemiluminescence immunoassay. Following analysis two SARS-COV-2 IgG positive and two IgG negative sera were used for the development of this assay.

### Assay platform preparation

A 96-well flat-bottom immunoplate (Extra Gene, USA) was coated with 100 µl/well of SARS-CoV-2 specific recombinant nucleoprotein (0.125 µg/well) in coating buffer (sodium bi-carbonate, pH >9) and incubated either at 37 °C for an hour or overnight at 4 °C. The unbound antigen was then decanted followed by blocking with 100 µl/well of blocking buffer (PBS, 0.1% Tween-20, 2% BSA) and incubated at 37 °C for an hour. Following incubation wells were washed three times with ELISA wash buffer (50 mm Tris, 0.05% Tween 20, 0.1% SDS, 0.8% NaCl, distilled water) and used for the assay.

### Assay Procedure

100μl of test serum at 1:100 dilution in diluent buffer (PBS, 0.1% Tween20, 1% BSA) was added into each well and incubated at 37 °C for 15 minutes. One positive, two negative and two plate controls (no serum was added) were added in each plate. After incubation, the contents of the wells were aspirated and the plate was washed 5 times using ELISA wash buffer. 100µl of optimized goat anti-human IgG conjugated with HRP was added to each well and then incubated for 10 minutes at 37 °C. Following incubation, the plate was washed 5 times and 100μl TMB was added into each well and incubated for 5 minutes at room temperature. Further colour development, 100μl 1.5M Sulfuric Acid (H_2_SO_4_) was used as stop solution and the optical density (OD) was measured at 450 nm using a Multiplex micro plate ELISA reader.

### Standardization and optimization of ELISA procedure

The procedure was optimized for antigen, conjugate, TMB and stop solution used in this study by checkerboard titration using the positive and negative control sera at different dilutions. The positive and negative sera were diluted at 1:50, 1:100 and 1:200 and tested against different concentrations of conjugate (dilutions 1:2000, 1:3000, 1:4000 and 1:5000) and coating antigen (dilutions 1:50, 1:100, 1:200 and 1:400). From the multiple combinations, the optimum condition that showed the best signal to noise ratio (S/N) with acceptable background has been selected. The formula used for calculating the S/N was,

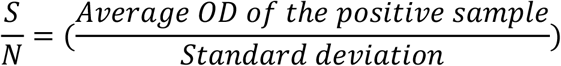

### Determination of the cut-off value

The cut off value has been determined with the negative samples. The mean OD of the negative controls has been determined and a sample is considered positive when the sample OD value at 450 nm exceeds the mean OD value of negative controls plus three times standard deviation (SD) defined as cut-off value. For a sample to be negative the value should be equal or less than the cut-off OD.

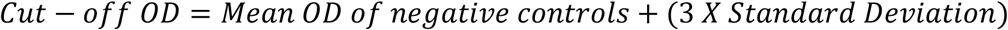

### In-house (Laboratory) performance evaluation

#### Reproducibility

Intra-assay variation was determined by providing 05 replicates of positive controls and negative controls in the same plate within a day. Inter-assay repeatability was checked by testing positive and negative controls at 15 different work days and coefficient of variation was determined using the following formula,

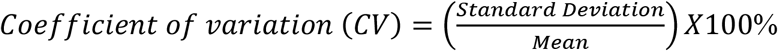

#### Clinical Validation

For clinical validation, sera from panel 1, 2 and 3 were assayed to determine clinical sensitivity and specificity of the assay.

### Statistical analysis

Panel 1 and 2 were used to determine sensitivity and panel 3 for assessing specificity and cross reactivity. Sensitivity, specificity, positive predictive value (PPV) and negative predictive value (NPV) and area under curve (AOC) with 95% confidence interval were estimated to see the effectiveness of this ELISA assay. Cohen’s Kappa test was used to evaluated the test agreement. Analysis was performed with STATA 13 (StataCorp, LP, College Station, Texas, USA) and GraphPad Prism 7.05 were used for graphical presentation.

## Results

### Characterization of seropositive and seronegative samples by Commercial Kit

Thirteen and seventeen serum samples from the panel 2 and panel 3, respectively, were tested by Elecsys Anti-SARS-CoV-2 immunoassay to determine and select sero-status as well as positive and negative controls. The result calculated by the analyser showed that all tested RT-PCR positive samples were reactive and the negative samples were non-reactive. Two positive controls (P1, P2) and two-negative controls (N1, N2) were selected for the development of this in-house rapid ELISA.

### Optimization of Assay protocol

Highest signal to noise ratio for positive controls with acceptable background from the checkerboard titration has been selected (Table 1). The optimal conditions for this ELISA include coating well with recombinant nucleocapsid antigen (NCP) at dilution 1:200, blocking for 1hr at 37 °C, sample dilution of 1:100 and detection anti-human IgG antibody-conjugate at 1:4000 dilutions, render the best possible result. The total assay time required to perform this assay was established 30 minutes.

**Table 1:** Checkerboard titration using positive samples.

|  | Conjugate ratio | NCP 1:50 | NCP 1:100 | NCP 1:200 | NCP 1:400 |
| --- | --- | --- | --- | --- | --- |
| Signal to noise | Conjugate 1:2000 | 9.920 | 12.60 | 13.23 | 11.80 |
| ratio (Average | Conjugate 1:3000 | 13.09 | 14.52 | 14.44 | 12.79 |
| positive/Standard | Conjugate 1:4000 | 13.02 | 14.13 | 13.56<sup>a</sup> | 10.69 |
| division) | Conjugate 1:5000 | 10.89 | 12.44 | 10.97 | 9.03 |

<sup>a</sup> represent the suitable S/N ratio for this study at sample dilution of 1:100

### Performance evaluation

#### Reproducibility and precision

As mentioned before, the intra-assay and inter-assay variation have been evaluated, with 05 replicates of positive and negative controls on the same day in a plate for the former, and testing the controls in 15 different days for the later. Coefficient of variation (CV) depicts reproducibility and precision and analysis showed CV was <25% for the controls used in inter assay and <10% for intra assay (Table 2).

**Table 2:** Reproducibility and Precision of the in-house ELISA.

| Controls used | CV (inter assay) | CV (intra assay) |
| --- | --- | --- |
| Positive control P1 | 16.46 | 5.04 |
| Positive control P2 | 19.33 | 7.61 |
| Negative control N1 | 21.59 | 4.964 |
| Negative control N2 | 20.48 | 9.454 |

#### Clinical Validation

The performance validation of the assay was conducted using serum samples collected from nineteen (n-19) RT-PCR confirmed positive patients within 14 days of onset of clinical symptoms, while sixty sera collected from COVID-19 RT-PCR cases at their convalescence stages (>14 days of onset of clinical symptoms). Total one hundred and five (n=105) sera were used as negative samples of which 81 samples from healthy donors and 24 samples from pre-pandemic dengue positive cases.

In patients, whose samples are collected within 14 days of onset of symptoms, among 19 RT-PCR confirmed positive patients, 16 showed positive IgG antibody titres to NCP with sensitivity of 84.2% (95% confidence interval, 60.4%, 96.6%) (Table 3). Within 14 days the test agreement between NCP ELISA and gold standard was found 81.4% (Kappa=0.814, p<0.001) (Table 3 and Fig 1A), with positive predictive value (PPV) 97.1% and negative predictive value (NPV) 84.2% (Table 4). While following the patients, seropositivity increased at day >14 and among 60 RT-PCR confirmed positive patients 59 cases were detected with sensitivity 98.3% (95% CI, 91.1%, 100.0%), whereas the test agreement was 94.8% (Kappa=0.948, p<0.001) (Table 3 and Fig 1A), with PPV and NPV 99.0% and 95.2%, respectively (Table 4). The overall test sensitivity is 94.9% (95% CI, 87.5%, 98.6%) (Table-3 and Fig 1B), with test agreement 92.2% (Kappa=0.922, p<0.001) and 96.2% PPV and NPV (Table 3 and 4). To check the specificity and cross reactivity we have run 81 sera from healthy donors and 24 from dengue positive samples. Among them only 3 samples were misdiagnosed and the overall specificity was 97.1% (95% CI, 91.9%, 99.4%) (Table 3 and Fig 1A).

**Table 3:** Specificity and Sensitivity analyses for NCP antigens against IgG in symptomatic and real time RT-PCR positive patients

| Days | AUC (95% CI) | Sensitivity, % (95% CI) | Specificity (95% CI) | Kappa | p-value |
| --- | --- | --- | --- | --- | --- |
| <14 | 0.91(0.82, 0.99) | 84.2(60.4, 96.6) | 97.1(91.9, 99.4) | 0.814 | <0.001 |
| >14 | 0.98(0.96, 1.00) | 98.3(91.1, 100.0) | 97.1(91.9, 99.4) | 0.948 | <0.001 |
| Overall | 0.96(0.93, 0.99) | 94.9(87.5, 98.6) | 97.1(91.9, 99.4) | 0.922 | <0.001 |

Note: AUC: Area under curve; 95% CI: 95% Confidence interval
Cohen's Kappa test was used to evaluate the test agreement

**Table 4.**
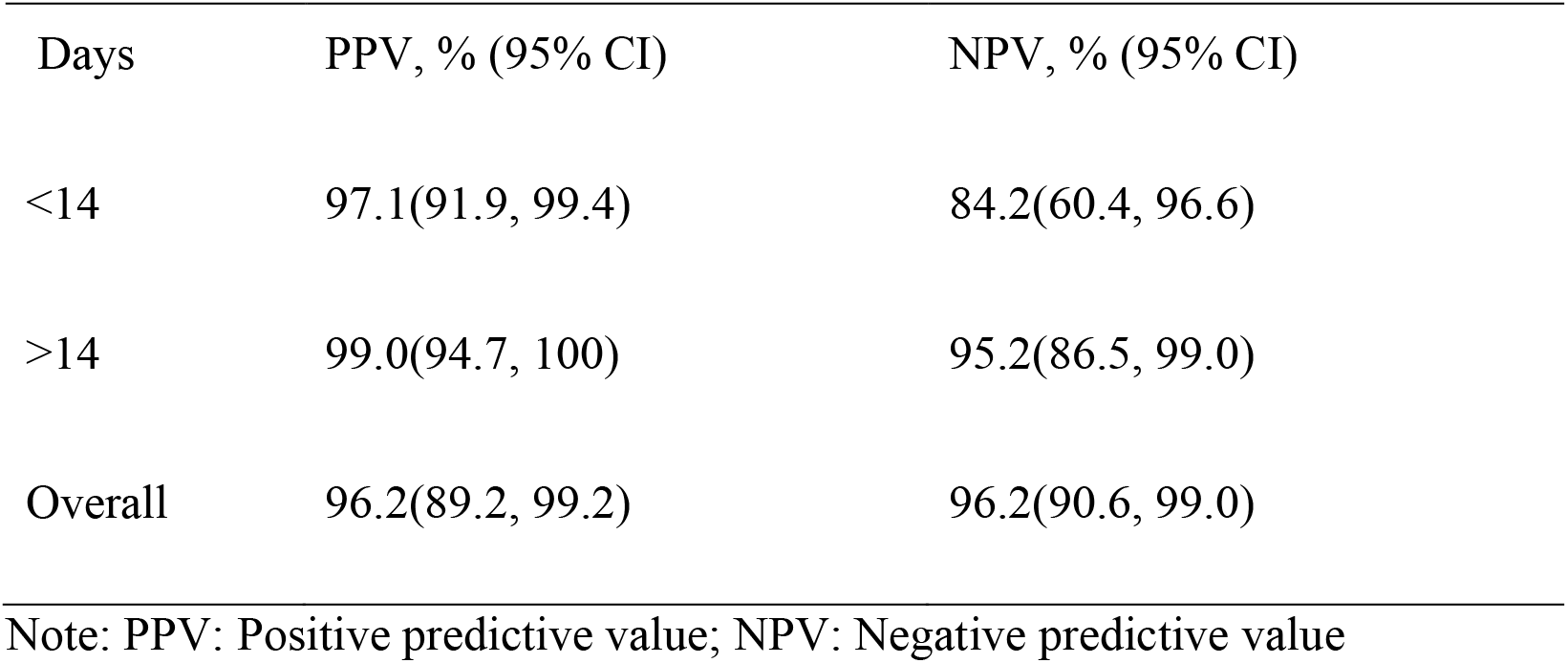
Positive and negative predicted value and test agreement of the assay procedure.

| Days | PPV, % (95% CI) | NPV, % (95% CI) |
| --- | --- | --- |
| <14 | 97.1(91.9, 99.4) | 84.2(60.4, 96.6) |
| >14 | 99.0(94.7, 100) | 95.2(86.5, 99.0) |
| Overall | 96.2(89.2, 99.2) | 96.2(90.6, 99.0) |

Note: PPV: Positive predictive value; NPV: Negative predictive value

**Fig 1.**
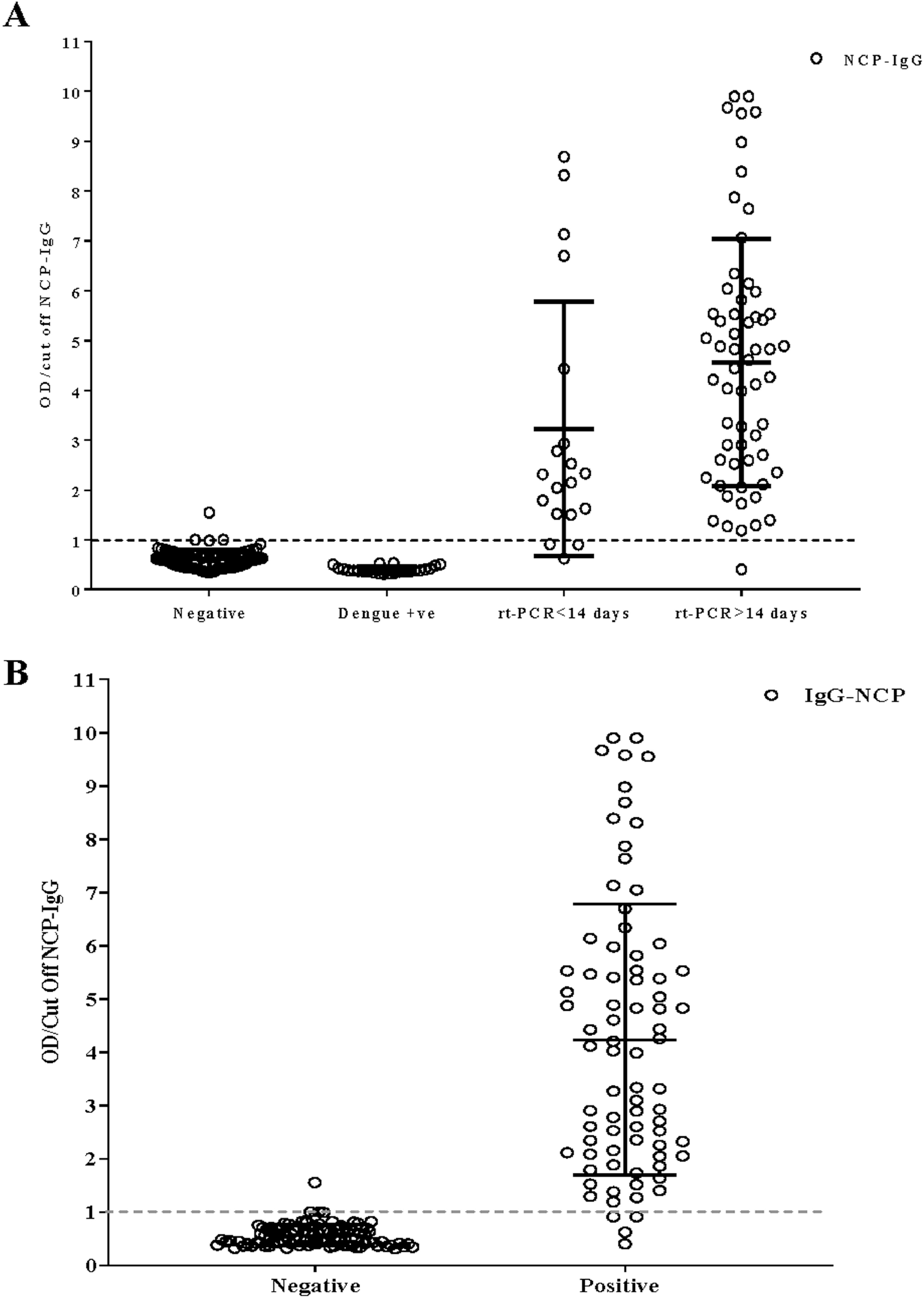
Detection of SARS-CoV2 nucleocapsid IgG among healthy donors, dengue positive samples, and SARS-CoV2 confirmed patients. Ratio of OD/cut off NCP-IgG value of negative, dengue positive and positive with SARS-CoV-2 (A) and all negative and positive cases (B) were shown. Data are presented as mean with ± Standard deviation. The reference line indicating the cut off of the in-house ELISA methods.

### Evaluation of NCP IgG ELISA with FDA approved commercial antibody immunoassay

Thirteen RT-PCR positive samples as well as seventeen negative samples have been tested at Immunobiology, Nutrition and Toxicology lab, Infectious Diseases Division, icddr,b. Both the sensitivity and specificity showed 100% for in-house ELISA while comparing with the FDA approved commercial antibody immunoassay (Table 5). The test agreement between Elecsys and in-house ELISA was 100%.

**Table 5:**
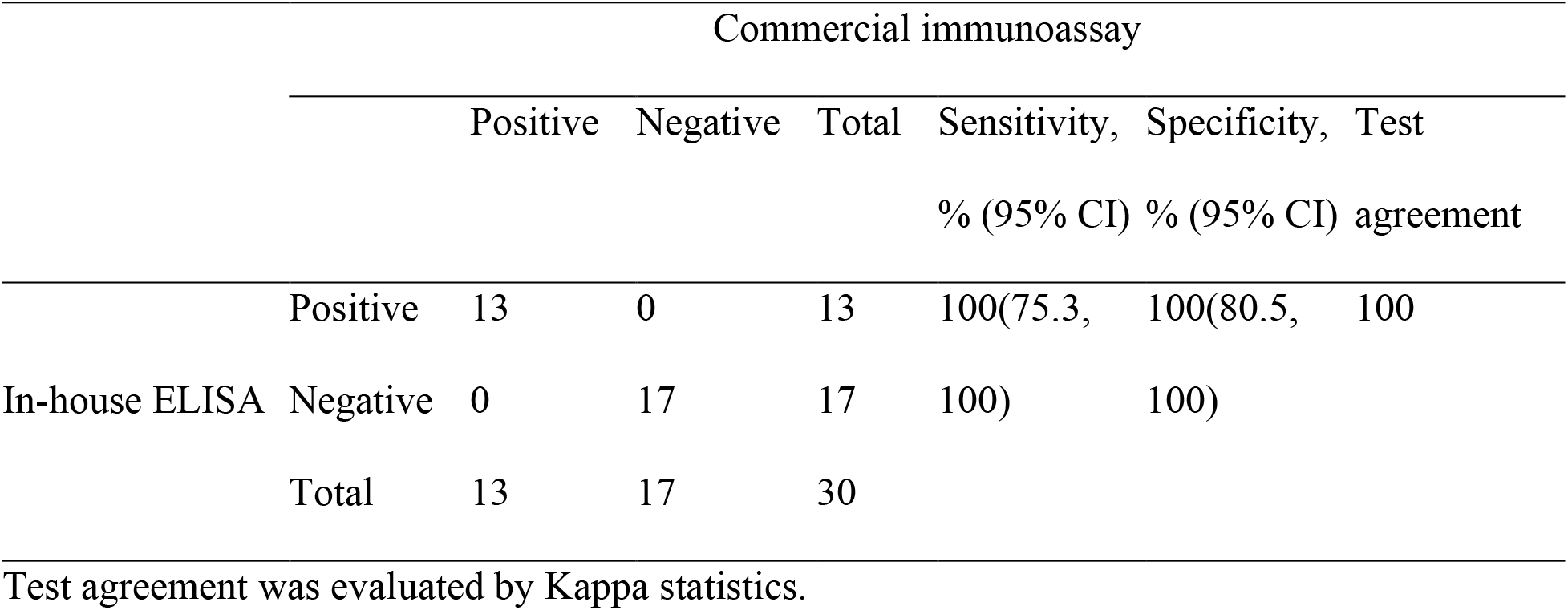
Comparison between in-house IgG ELISA with FDA approved commercial antibody immunoassay.

|  |  | Commercial immunoassay |  |  |  |  |  |
| --- | --- | --- | --- | --- | --- | --- | --- |
|  |  | Positive | Negative | Total | Sensitivity,<br>% (95% CI) | Specificity,<br>% (95% CI) | Test<br>agreement |
| In-house ELISA | Positive | 13 | 0 | 13 | 100(75.3, | 100(80.5, | 100 |
|  | Negative | 0 | 17 | 17 | 100) | 100) |  |
|  | Total | 13 | 17 | 30 |  |  |  |

Test agreement was evaluated by Kappa statistics.

## Discussion

On the context of the current pandemic situation, several serological tests based on either chemiluminescence, lateral flow, neutralization or immunosorbent assay have been developed and approved by FDA for emergency use (19, 22, 23). Eleven of these 58 serological tests are mounted on the enzyme linked immunosorbent assay targeting either IgM, IgG or total antibody (19).

The biologics, biosimilars, and bio-diagnostics developed by different biopharmaceuticals and biosimilar manufacturer to combat COVID-19, fall short to satisfy the global demand surpassing in-land need. Different countries are thus inclined to manufacturing their own diagnostics to satisfy national demand (24-26). Late-entrant countries in biopharmaceutical industry, like Bangladesh, are now working towards developing their own biosimilar products, anticipating the forthcoming situations when these countries will have to lose access to World Trade Organisation (WTO) waiver as a consequence of leaving LDC category in 2024 (27-31). Bangladesh, with death toll of 6388 and a total of 447,341 infected cases (till 22 November, 2020) (32) has yet to implement any immunoassay kit for the management of COVID-19 that meet the standard set by its drug authority. Although, policy makers are giving emphasis on the implementation of proper immunoassay kits, in-house assay kits to meet the demand are essential to manage the wreckage. The emerging condition underpins our endeavour to develop an indigenous IgG-ELISA specific to COVID-19, an approach to creating an opportunity to satisfy national and global demand.

This assay is mounted upon nucleocapsid as an antigen which provides increased sensitivity compared to either Spike-1 (S1) or Receptor Binding Domain (RBD) for detecting early phase of infection due to its primordial inception (21, 33, 34). High expression of NCP protein of coronaviruses has already been reported during infection (35), which is not only B-cell immunogen but also evoke cellular immune response in SARS infected patients (36, 37). Also, spike (S) gene of SARS-CoV-2 has 76% similarities with that of SARS-CoV-1, which exhibits non-synonymous mutations as the disease evolves over time (38-40). On the other hand, the nucleocapsid protein is more conserved having 90% amino acid homology with SARS-CoV-1 (39) which affected South-East Asian countries comparatively to a lesser extent (41).

For tropical and sub-tropical dengue-endemic countries (42) developing serological tests specific to COVID-19 is quite challenging, for serological and symptomatological overlap between the diseases in question (43, 44). Misdiagnosis of COVID-19 as dengue due to serological cross-reactivity has already been reported in Indonesia (45, 46). Singapore (47) and Thailand (48) and vice-versa for rapid COVID-19 antibody kits in India (49). To address the concern, our assay is characterized including 24 dengue positive samples from pre-COVID-19 situation that are found to be non-cross-reactive in our in-house IgG ELISA.

This assay system exhibited 100% sensitivity as well as specificity in relation with the commercial kit. A total of 184 samples subsumed in the panels have been then assayed by in-house ELISA and our assay exhibits a sensitivity of 84% for samples that have been collected within 14 days of symptom onset, and reach to 98% for samples collected after 14 days. This finding is in accordance with Long et al and others where the seroconversion for IgG peaked at 100% within 17-19 days of onset of symptoms (50). Our study suggests the use of this in-house ELISA in early phase of infection detection as well as in retrospective serosurvey.

Diagnostic settings, handling a surge of samples, require a time-saving, convenient test method. The strength of our indigenous system lies here, assay time being optimized at total 30 minutes, while currently available ELISA kits such as “Euroimmun Anti-SARS-CoV-2 ELISA (IgG)” or “BioRad Platelia SARS-CoV-2 Total Ab” require total of 120 minutes of incubation to perform their assay (19). Spectrum bias, which may affect sensitivity calculation, is circumvented by longitudinal antibody analysis of individuals whose sera have been exploited as positive controls. (51). Also the positive panels comprises of multiple samples from three patients who exhibited higher antibody titer for a long period which is mentioned in a previous study (52).

Certain limitations in our assay development exist that are to be addressed. Firstly, for cross reactivity test, no known respiratory sample was assessed, and secondly, the cohort sample size was actually inadequate to draw conclusions on samples collected within 0-14 days of symptom onset.

In conclusion, this in-house ELISA demonstrates its usefulness for the early detection as well as for serosourveillence of SARS-CoV-2 IgG that developed against nucleocapsid proteins. This assay is easy to perform, cost-effective and results can be interpreted at 30 minutes of test run. Moreover, this test showed comparable level of performance against commercial FDA approved electrochemiluminescence immunoassay and detected IgG from SARS-CoV-2 infected patients. Hence, this SARS-CoV-2 NCP-IgG Rapid ELISA could be equally applied as one of the COVID-19 early sero-monitoring tools all over the COVID-19 pandemic countries.

## Data Availability

All data underlying the findings in our study are freely available in the manuscript. For additional information please refer to: http://www.grblbd.com

## Acknowledgement

The authors express their thanks all the participants who were willing participate in the study. They also thank their team members for their support work during the current pandemic situation.

## Author Contribution

Nihad Adnan and Bijon Kumar S. designed and supervised this research. Mohd. Raeed J., Mumtarin Jannat O., Nowshin Jahan., Shahad Sharif K., Tammana Ali. performed the experiment. Md. Ahsanul H. performed the data curation and statistical analysis. Mohib Ullah K., Eiry Kobatake, and Masayasu Mie proof read it. All authors contributed to the construction and writing this article.

## Ethical Conduct of Research

This study was approved by the Bangladesh Medical Research Council (BMRC)

